# Evaluating Genomic Polygenic Risk Scores for Childhood Acute Lymphoblastic Leukemia in Latinos

**DOI:** 10.1101/2023.06.08.23291167

**Authors:** Soyoung Jeon, Ying Chu Lo, Libby M. Morimoto, Catherine Metayer, Xiaomei Ma, Joseph L. Wiemels, Adam J. de Smith, Charleston W.K. Chiang

## Abstract

The utility of polygenic risk score (PRS) models has not been comprehensively evaluated for childhood acute lymphoblastic leukemia (ALL), the most common type of cancer in children. Previous PRS models for ALL were based on significant loci observed in genome-wide association studies (GWAS), even though genomic PRS models have been shown to improve prediction performance for a number of complex diseases. In the United States, Latino (LAT) children have the highest risk of ALL, but the transferability of PRS models to LAT children has not been studied. In this study we constructed and evaluated genomic PRS models based on either non-Latino white (NLW) GWAS or a multi-ancestry GWAS. We found that the best PRS models performed similarly between held-out NLW and LAT samples (PseudoR^2^ = 0.086 ± 0.023 in NLW vs. 0.060 ± 0.020 in LAT), and can be improved for LAT if we performed GWAS in LAT-only (PseudoR^2^ = 0.116 ± 0.026) or multi-ancestry samples (PseudoR^2^ = 0.131 ± 0.025). However, the best genomic models currently do not have better prediction accuracy than a conventional model using all known ALL-associated loci in the literature (PseudoR^2^ = 0.166 ± 0.025), which includes loci from GWAS populations that we could not access to train genomic PRS models. Our results suggest that larger and more inclusive GWAS may be needed for genomic PRS to be useful for ALL. Moreover, the comparable performance between populations may suggest a more oligo-genic architecture for ALL, where some large effect loci may be shared between populations. Future PRS models that move away from the infinite causal loci assumption may further improve PRS for ALL.

## INTRODUCTION

Acute lymphoblastic leukemia (ALL) is the most common type of childhood cancer worldwide, representing 20% of all cancers in children in the United States.^1^ There are few established environmental risk factors for ALL, and genome-wide association studies (GWAS) have confirmed the contribution of genetic variation to ALL risk. To date, at least 19 loci have been discovered and replicated in previous GWAS, primarily performed with European ancestry individuals, suggesting the polygenic nature of susceptibility to ALL^2–12^. Yet, how these variants collectively contribute to disease risk has not been fully characterized.

Polygenic Risk Scores (PRS) can identify individuals at significantly elevated risk for a disease, such as cancer, by providing a quantitative measure of an individual’s inherited risk based on the cumulative impact of variants shown to be associated with the disease of interest. Moreover, there has been growing evidence that the predictive power of PRS can be further increased by aggregation of genotypic effects across all variants even if they do not reach the commonly acknowledged genome-wide significance threshold for association (*P* = 5e-8).^13,14^ With ALL, this genomic PRS approach may enhance the efficacy of PRS models given the small number of known susceptibility loci.

However, one of the biggest limitations of PRS is the lower predictive performance in non-European ancestry populations.^15^ Part of this loss in efficacy may be due to the over-representation of GWAS participants of European ancestry,^15,16^ resulting in much more informative GWAS for European ancestry individuals compared to that for other ancestries. The poor transferability may also arise due to differences between patterns of linkage disequilibrium (LD), allele frequencies, causal variants, and effect sizes.^15,17,18^ Such a limitation is particularly important for ALL, since Latino children have an higher and faster-increasing risk and poorer survival than non-Latino Whites in the United States.^19–23^ Currently available PRS models for ALL are based only on a limited number of known risk alleles. One of the first PRS models for ALL was one constructed with 11 single nucleotide polymorphisms (SNPs) known to be associated with ALL as of 2018, with effect sizes estimated from a European ancestry cohort.^24^ Its efficacy in individual risk discrimination analysis may be over-estimated as early estimates of variant effect sizes tend to be over-estimated, and its transferability to non-European cohorts has not been evaluated. A subsequent PRS model reported in 2021 using again only SNPs from known associated loci from multi-ancestry GWAS showed lower predictive performance than the earlier study, though it demostrated similar performance between Latinos and non-Latino White cohorts.^2^ No study has constructed genomic PRS models for ALL in any population to date.

In this study, we set out to construct and evaluate genomic PRS models derived using non-Latino White (NLW) cohorts and test their transferability to Latino (LAT) individuals. We evaluated two genomic PRS approaches – Pruning and Thresholding (P+T) and LDPred2 – in parallel to PRS models constructed based on only genome-wide significant loci from the literature. We also aimed to examine whether effect sizes estimated from ethnic-specific GWAS or multi-ancestry meta-analysis, and whether training with matched ancestry LD reference panel, could improve the efficacy of the PRS.

## RESULTS

For each step of, we used three non-overlapping datasets to (1) perform discovery GWAS to estimate variant effect sizes, (2) optimize parameters for the best predictive score, and (3) evaluate the predictive performance of the resulting scores (**Figure 1**). Following the convention previously suggested,^25,26^ we refer to the datasets used in each of the three steps as “GWAS”, “testing”, and “validation” datasets. We randomly selected and held out 360 cases and 1,200 controls from each of NLW and LAT as the testing datasets to identify the best PRS models, and reserved the entire California Childhood Leukemia Study (CCLS) cohort for validation. We used the remaining samples as the GWAS dataset in the three different discovery GWAS: (1) NLW-only meta-analysis, (2) LAT-only GWAS, or (3) multi-ancestry meta-analysis. For each GWAS, we constructed PRS using two established approaches - Pruning and Thresholding (P+T) and LDPred2^27^. We explored multiple strategies to develop PRS models. We labelled the different strategies using the convention of “POP_GWAS__POP_testing_”, where POP_GWAS_ is the population in which the discovery GWAS was conducted, and POP_testing_ is the population in which the optimization for the best model was performed (**Methods**). Once the best performing PRS model for a given strategy is identified, the PRS model was then evaluated in the validation cohort - CCLS NLW (306 cases and 258 controls) or LAT (592 cases and 509 controls).

**Figure 1:**
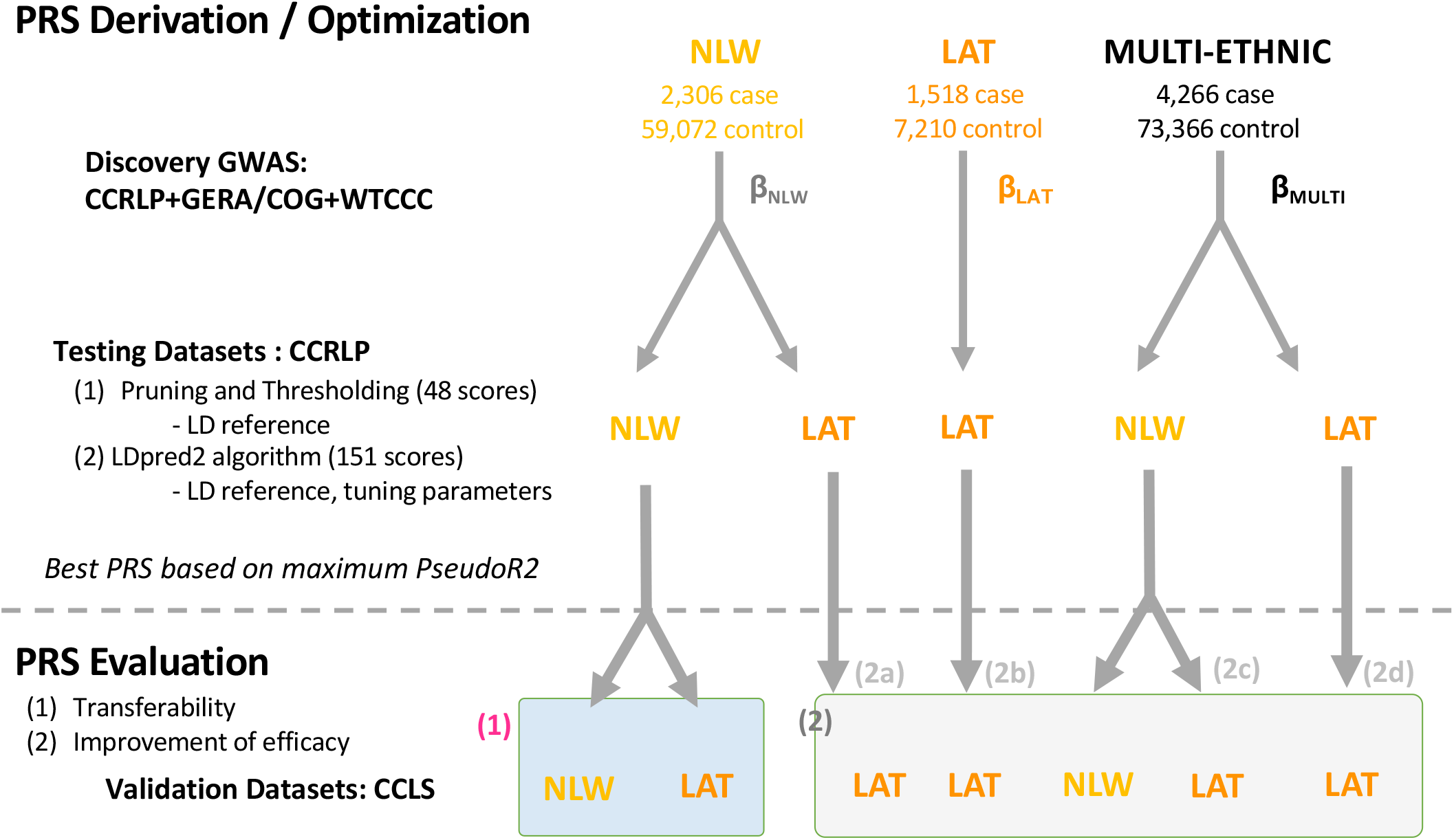
summary of study design and analysis. The flowchart details different cohorts used for each step of PRS derivation with different discovery GWAS dataset, optimization dataset, and evaluation in either non-Latino white or Latino populations. In PRS evaluation, comparison (1) focused on evaluating the transferability of PRS models optimized in non-Latino White cohort. Comparison (2) focused on different strategies for improving the PRS efficacy by optimizing in a Latino cohort (2a), using a Latino-only discovery GWAS (2b), using a multi-ethnic discovery GWAS and optimized in non-Latino white (2c) or Latinos (2d). NLW: no-Latino White, LAT: Latino American, CCRLP: California Childhood Cancer Record Linkage Project, GERA: Genetic Epidemiology Research on Aging Cohort, COG: Children’s Oncology Group, WTCCC: Wellcome Trust Case-Control Consortium, CCLS: California Childhood Leukemia Study.

### Transferability of genomic PRS for ALL

We first evaluated a genomic PRS model derived from GWAS summary statistics of a NLW cohort and its transferability to the LAT cohort. We performed a GWAS in 2,306 cases and 59,072 controls in NLW (**Supplemental Table 1**) after holding out individuals for testing and validating the PRS models. Our first design is termed NLW_NLW, for the discovery GWAS was performed in NLW, and the model was optimized also in held-out NLW samples (strategy 1, **Figure 1**). This is a typical scenario where GWAS and PRS model optimizations were both completed in European-ancestry populations.

The best model with the highest Negelkerke’s Pseudo R^2^ in the NLW_NLW approach was based on LDPred2, a non-sparse model with ρ = 0.0032 and *h*^2^ = 0.22. This model consisted of approximately 1.08M SNPs across the genome and is significantly associated with case/control status in both CCLS NLW and LAT cohorts (*P* = 4.1e-9 and 3.9e-12 for NLW and LAT, respectively). The resulting PRS explained 8.6 ± 3.2% of the variance in the CCLS NLW cohort, after accounting for covariates, as measured by pseudo R^2^ (**Table 1**). The same PRS model explained 6.0 ± 2.0% of the variance in the CCLS LAT cohort (**Table 1, Figure 2**), suggesting minimal loss of transferability in efficacy after taking into account the standard errors of these estimates. The AUC in both NLW and LAT are also similar (0.667 ± 0.045 and 0.652 ± 0.032 in NLW and LAT, respectively, in the full prediction model, including PRS as well as sex and 10 principal components; **Table 1**).

**Table 1:**
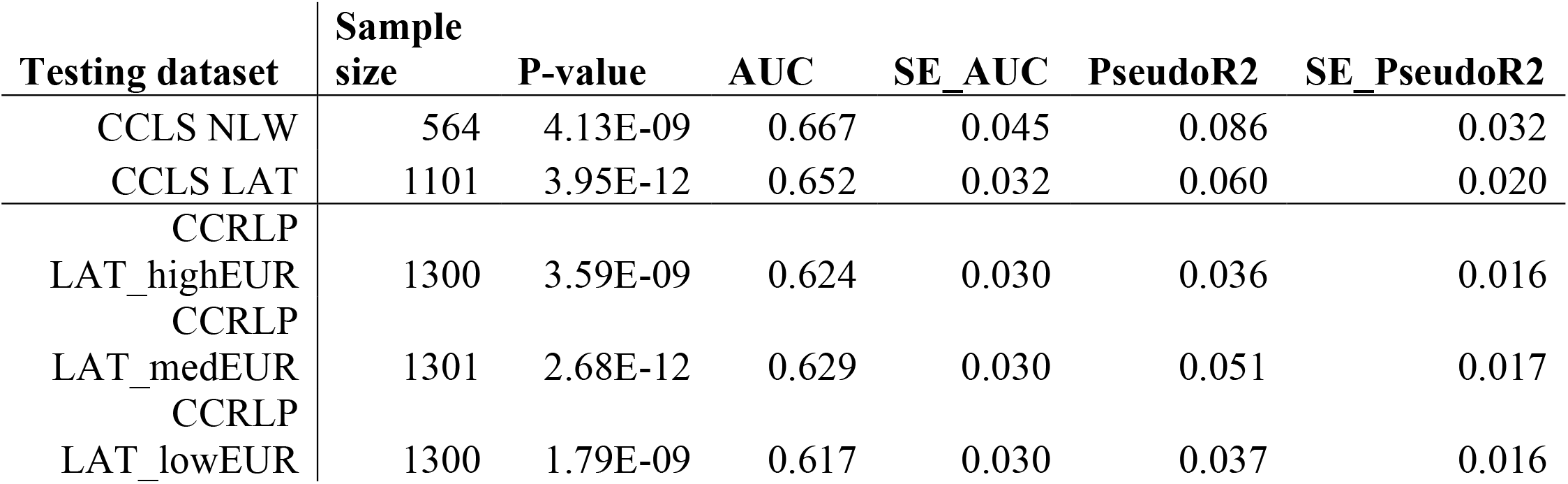
Performance of the best model for NLW_NLW strategy across different testing datasets. P-value denotes the evidence of association of the PRS model in a logistic regression model with additional covariates of 20 PCs and sex. AUC denotes area under the curve from receiveer-operator characteristic analysis. PseudoR2 was calculated from the difference between a logistic regression model with PRS and one without PRS. SE denotes standard error for both AUC and PseudoR2, which were computed using 1,000 bootstrap samples. CCRLP LAT_highEUR, medEUR, and lowEUR denote the top, middle, and bottom tertile, respectively, of individuals sorted by proportion of estimated European ancestries.

**Figure 2:**
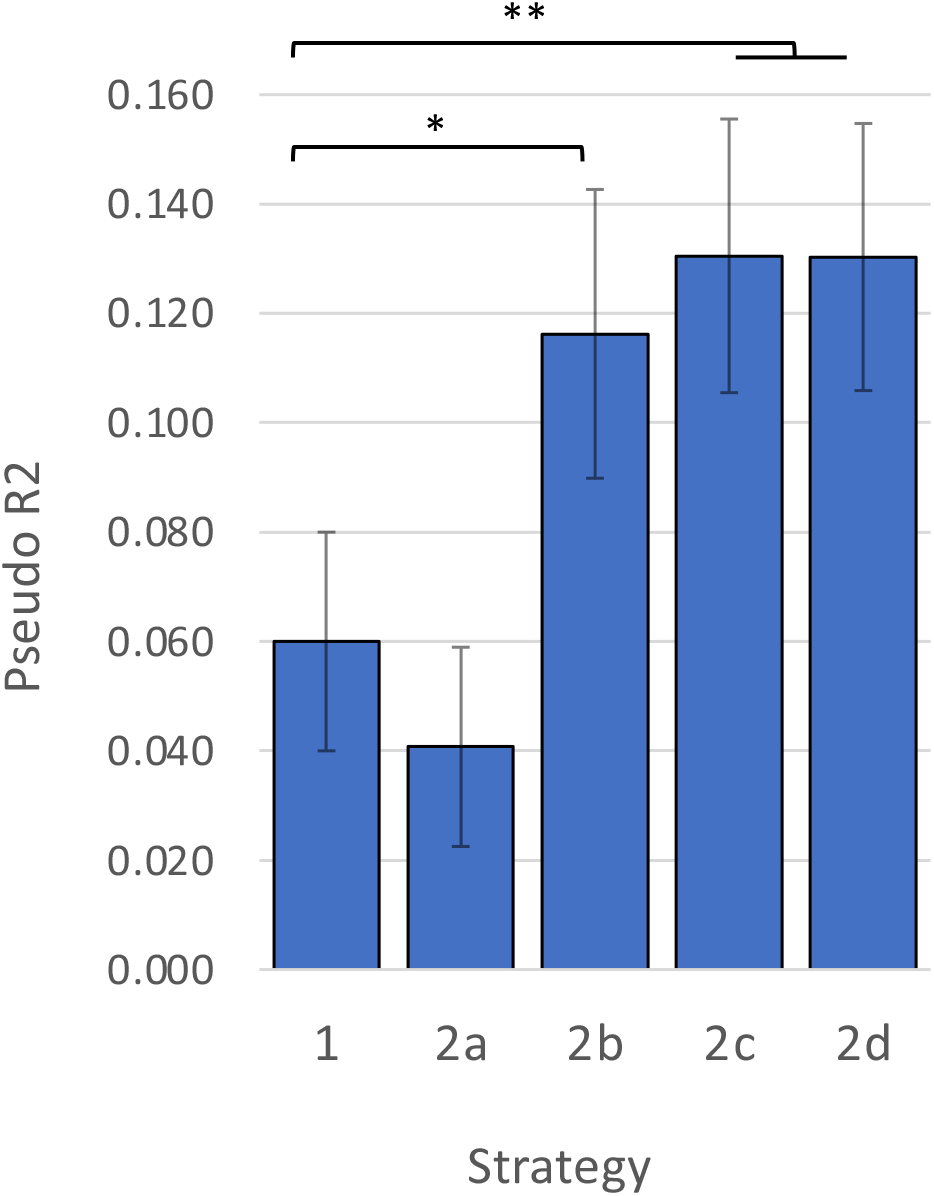
PRS efficacy based on the best performing model for each strategy, as validated in CCLS Latinos. The PRS efficacy, as measured by pseudo R2, of the best performing model for each strategy aimed to improve the efficacy of PRS models for LAT (strategies 2a-2d in Figure 1) is summarized and compared to a baseline model (strategy 1). Baseline model is a Euro-centric discovery GWAS (using only NLW individuals), with model optimization in held-out NLW individuals. * Strategy 2b (LAT_LAT; Latino-only discovery GWAS and PRS model optimization in LAT) is significantly better than the baseline model (*P* = 0.0019). ** Strategy 2c and 2d (META_NLW and META_LAT; Multi-ancestry meta-analysis discovery GWAS and model optimization in NLW or LAT, respectively) are both better than the baseline model as well (*P* < 1e-4). Statistical significance is computed by 10,000 rounds of bootstrap samples across individuals. No other comparisons produced statistically significant results.

An alternative approach to evaluate the transferability of the genomic PRS model is to assess if the prediction efficacy differs by proportion of European ancestry in the Latino individuals. Because the CCRLP has the largest collection of LAT individuals and have not been used in the NLW_NLW model, we can evaluate the prediction accuracy in CCRLP LAT individuals (N = 3901; 1878 cases). In tertiles of LAT individuals, each with approximately 1300 individuals, we found little evidence of differences in performance across strata of ancestry proportions (Pseudo R^2^ = 0.036, 0.051, 0.037 across the highest, middle, and lowest tertiles by European ancestries in LAT; AUC = 0.624, 0.629, and 0.617, respectively. **Table 1**). Taken together, we identified little evidence that there is a substantial difference in transferability between NLW and LAT populations or ancestries.

### Improving the prediction accuracy of genomic PRS for Latinos

We first evaluated a scenario where the LAT was used as the cohort to identify the optimal PRS model, even though the discovery GWAS was still from NLW (NLW_LAT; strategy 2a in **Figure 1**). We found that in this case, the best model was a LDPred2 sparse model with parameters ρ= 0.01 and *h*^2^ = 0.1826. This model did not appear to improve the performance of the PRS in CCLS LAT over the best NLW_NLW model (PseudoR^2^ = 0.041 ± 0.018, compared to 0.060 ± 0.020 under NLW_NLW approach; **Figure 2, Supplemental Table 2**).

We also evaluated a scenario where the LAT were used both for the discovery GWAS and PRS model optimization. In this case, 1,518 cases and 7,210 controls of LAT individuals from CCRLP+GERA were used in the discovery GWAS, and 151 models based on either P+T or LDPred2 were optimized in 360 cases and 1200 controls from held-out LAT individuals (LAT_LAT strategy; 2b in **Figure 1**). The best PRS model from this approach was a LDPred2 sparse model with parameters ρ= 0.001 and *h*^2^ = 0.1764. When validating this model in CCLS, the performance was significantly better than when the NLW had been used for discovery GWAS (Pseudo R^2^ = 0.116 ± 0.026, compared to 0.060 ± 0.020 under the NLW_NLW strategy, *P* = 0.0019; **Figure 2; Supplemental Table 2**).

Finally, as discovery GWAS based in NLW or LAT are both potentially underpowered, we also evaluated the meta-analysis design that combined all four multi-ancestry cohorts from CCRLP+GERA as well as the COG+WTCCC samples (**Supplemental Table 1**). In total, the GWAS contained 4,226 cases (2306, 1518, 318, and 124 in NLW, LAT, EAS, and AFR, respectively) and 73,366 controls (59072, 7210, 5017, and 2067 in NLW, LAT, EAS, and AFR, respectively). We then trained the best genomic PRS model in either NLW (META_NLW; strategy 2c in **Figure 1**) or LAT (META_LAT; strategy 2d in **Figure 1**), both using a held-out sample of 360 cases and 1200 controls. Likely due to the increased sample sizes, the meta-analysis designs produced the best performing genomic PRS models. Under the META_NLW design, the best model was a LDPred2 sparse model with parameters ρ= 0.0032 and *h*^2^ = 0.1376. Under this model, the prediction accuracy in CCLS LAT was better than the naïve NLW_NLW strategy (Pseudo R^2^ = 0.131 ± 0.025 vs. 0.060 ± 0.020, *P* < 1e-4; **Figure 2, Supplemental Table 2**), and slightly though not significantly higher than the LAT_LAT strategy (Pseudo R^2^ = 0.116 ± 0.026; *P* = 0.15). The best META_LAT strategy (a non-sparse model with parameters ρ= 0.001 and *h*^2^ = 0.1127) also appeared to perform similarly compared to the META_NLW approach (Pseudo R^2^ = 0.130 ± 0.024; **Figure 2, Supplemental Table 2**). The AUC for the full model including PRS, sex, and PCs were 0.700 and 0.701 for META_NLW and META_LAT strategies, respectively. Our results thus suggest that given the currently available data, combining the largest multiethnic sample for discovery GWAS will lead to the best genomic PRS model in terms of prediction accuracy.

As the multi-ethnic meta-analysis GWAS is the most powerful discovery GWAS currently available, we also evaluated the transferability of the PRS model from the META_NLW strategy by comparing the PRS performance in CCLS NLW vs. LAT samples. The Pseudo R^2^ remains comparable between the two cohorts (e.g. under the META_NLW strategy, Pseudo R^2^ = 0.153 ± 0.034 for NLW vs. 0.131 ± 0.025 for LAT; **Figure 3, Supplemental Table 3**). This result is consistent with the attempt described above (NLW_NLW strategy).

**Figure 3:**
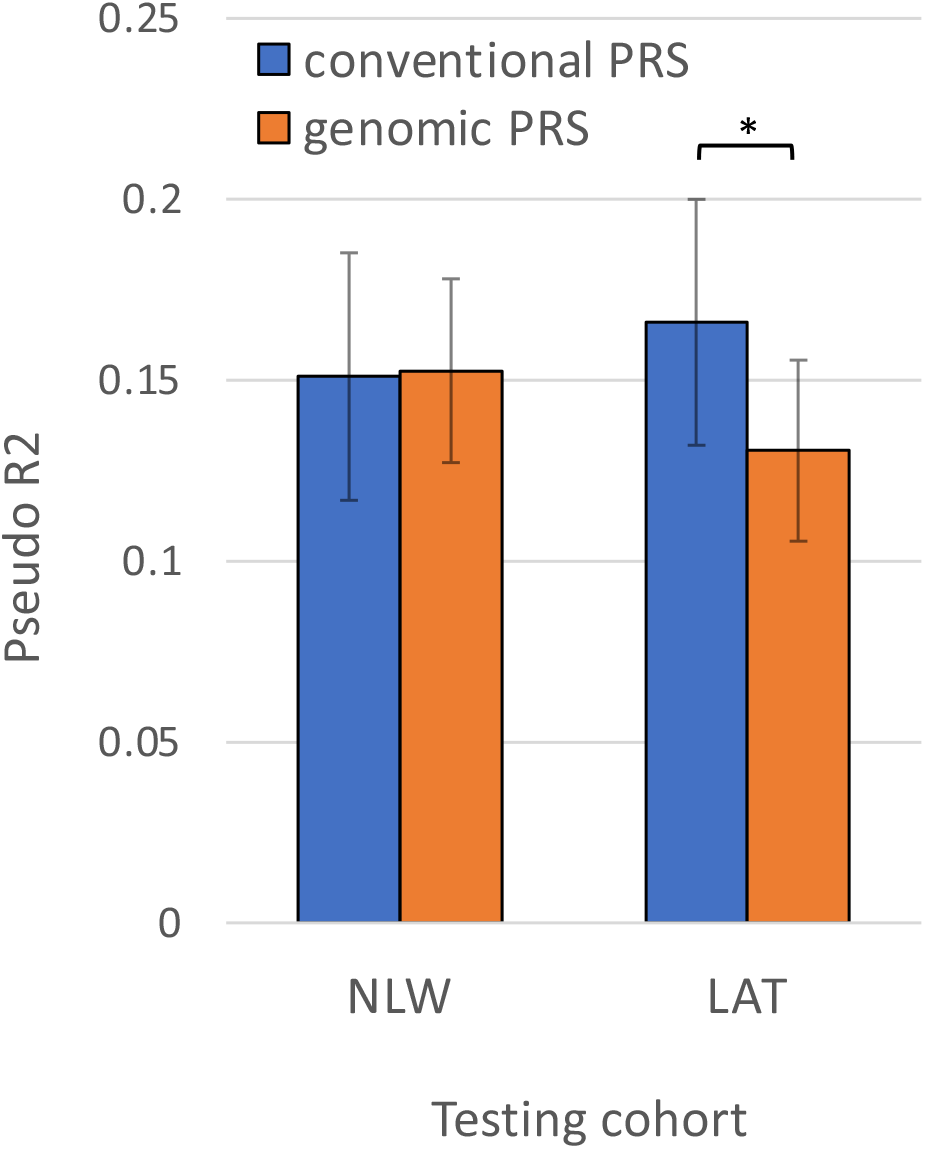
Predictive performance of best performing genomic PRS model (META_NLW) vs. PRS model constructed with 23 known ALL risk SNPs. Both models were tested in CCLS NLW and LAT, which were not used in the discovery GWAS for genomic PRS. CCLS was also not used in the identification of the 23 known loci in literature, although it had been used as replication cohort. * In CCLS LAT, the pseudo R2 for PRS model based on 23 known loci is significantly better than that from our best performing genomic PRS model (*P* < 1e-4) based on 10,000 sets of bootstrapping.

### Genetic Architecture of ALL

LDPred2 has two different modes of inference, LDPred2-grid and LDPred2-inf, where the former assumes some proportions of the variants are causal and parameters need to be optimized in a grid, while latter assumes an infinitesimal model where every variant have a mean effect of 0 with some small variance. In our META_NLW and META_LAT approaches, where we have the most powerful discovery GWAS to guide PRS model constructions, we noticed that LDPred2 models consistently outperformed the LDPred2-inf models (*e*.*g*. Pseudo R2 = 0.130 ± 0.025 in LDPred2 vs. 0.013 ± 0.016 in LDPred2-inf when model under the META_LAT strategy was evaluated in CCLS LAT; **Supplemental Figure 1, Supplemental Table 3**). Our results are thus consistent with a more oligogenic architecture of ALL, while LDPred2-inf is more appropriate for traits with highly polygenic inheritance.

### Genomic PRS vs. PRS based on genome-wide significant loci

Generally speaking, genomic PRS models, whether through P+T, LDPred2, or other similar approaches, are expected to be more accurate in risk prediction or stratification over a simple PRS model based solely on the set of known GWAS loci (i.e. those that have been shown to reach a P-value less than 5e-8 in one or more GWAS for a particular trait)^27^. Indeed, in each of the strategies that we have examined, the best genomic PRS models tend to be better than P+T model with p-value threshold of 5e-8, a special case which is equivalent to building a PRS model with the genome-wide significant loci. For instance, under the META_LAT strategy, the best PRS model achieved a pseudo R^2^ of 0.130 ± 0.024, while the best P+T model with P-value threshold of 5e-8 only attained a pseudo R^2^ of 0.088 ± 0.021.

However, the genomic PRS requires a held-out sample to optimize the parameters for building the PRS. This necessitates a reduction in the sample sizes available for GWAS. While this may not be a huge obstacle for common diseases, it could be a concern for a rare disease such as ALL. In order to evaluate the genomic PRS, we had to reduce our case proportions by 8.7% (from 2666 cases to 2306 cases and from 1878 cases to 1518 cases after removing 360 cases each for NLW and LAT respectively from CCRLP+Kaiser as training sample). Thus, an alternative approach could have been constructing a simple PRS model based on only genome-wide significant variants, and subsequently test this PRS model in independent cohorts.

We built a PRS model using 23 SNPs previously associated with ALL, identified across 11 studies^2–12^. These 23 SNPs were derived from 19 loci, including conditionally independent secondary associations at 4 loci (**Supplemental Table 4**). These associated SNPs were identified in one or more independent cohorts in the literature, including the full CCRLP+GERA datasets that were used for constructing and evaluating genomic PRS models above. Because there is no need to optimize the PRS model in held-out samples, we directly tested this “conventional” PRS in the independent CCLS cohort that were not used in the discovery of any of these 23 loci (although they had been used as part of the replication cohort in previous studies). This strategy produced better prediction accuracy than the best performing genomic PRS models in CCLS LAT (Pseudo R^2^ = 0.166 ± 0.025; AUC = 0.726 compared to Pseudo R^2^ = 0.131 ± 0.025 from genomic PRS derived using the META_NLW strategy, *P* < 1e-4; **Table 2, Figure 3**), a difference that was not seen between the conventional PRS and genomic PRS tested in CCLS NLW (Pseudo R^2^ = 0.151 ± 0.034; AUC = 0.706 compared to Pseudo R^2^ = 0.153 ± 0.034 from genomic PRS derived using the META_NLW, **Table 2, Figure 3**).

**Table 2:** Predictive performance of best performing genomic PRS model vs. conventional model constructed with 23 known ALL risk SNPs. Conventional PRS is a model based on 23 SNPs in literature known to be associated with ALL, having passing the genome-wide significance threshold of 5e-8 in GWAS. PseudoR2 was calculated from the difference between a logistic regression model with PRS and one without PRS. SE denotes standard error, estimated from 1000 bootstrap samples.

|  | CCLS NLW |  |  | CCLS LAT |  |  |
| --- | --- | --- | --- | --- | --- | --- |
|  | PseudoR2 | SE | AUC | PseudoR2 | SE | AUC |
| conventional PRS | 0.151 | 0.034 | 0.706 | 0.166 | 0.025 | 0.726 |
| genomic PRS | 0.153 | 0.034 | 0.710 | 0.131 | 0.025 | 0.700 |

## DISCUSSION

In the current study, we leveraged the largest available multi-ancestry meta-analysis GWAS to investigate strategies to build and evaluate polygenic risk score models for ALL across populations. We evaluated the extent of loss in efficacy for PRS models trained solely in NLW populations but applied in LAT populations, explored approaches to improve PRS models for LAT through different optimization strategies, and compared the genomic PRS models against a simple model that used all previously reported genome-wide significant ALL-associated variants with no optimization. We found little evidence of a loss in efficacy when transferring the genomic PRS model between populations. We also found that while leveraging multi-ethnic information to increase GWAS sample sizes and representation could lead to much more effective genomic PRS models (pseudo R^2^ = 0.131 ± 0.025, AUC = 0.700), this model currently still has lower prediction accuracy for Latinos compared to a simple model of using only 23 known ALL-associated SNPs (pseudo R^2^ = 0.166 ± 0.025, AUC = 0.726) that were derived from multiple independent cohorts in literature (including ones we do not have access to and not utilized in this study for building genomic PRS).

We undertook multiple analytical approaches to evaluate the transferability of a PRS model for ALL, but generally found little evidence of loss in efficacy across populations. After determining the best predictive PRS models in NLW, using either the CCRLP NLW (NLW_NLW approach) or the meta-analysis (META_NLW) for the discovery GWAS, we observed little difference in performance in CCLS NLW and LAT subjects (66.7% in NLW vs. 65.2% in LAT by AUC for the NLW_NLW strategy; 71.0% in NLW vs. 70.0% in LAT by AUC for the META_NLW strategy; **Table 1, Supplemental Table 3**). We also did not observe differences in the PRS predictive performance across strata of LAT individuals by estimated European ancestry proportions (**Table 1**). If there had been overt transferability issues, we would expect that strata with the highest European ancestry would have higher prediction accuracy compared to strata with lower European ancestry. It remains unclear whether the similar performance across populations is driven by representation in GWAS, European-ancestry admixture in LAT samples, or sufficiently shared genetic architecture between populations for ALL that is minimally impacted by LD differences.

As one possible explanation for comparable PRS efficacy between LAT and NLW is shared genetic architecture; whereby ALL may follow a more oligogenic architecture where several large effect causal loci exist on top of a polygenic background of smaller effect causal loci. Our previous study^2^ had demonstrated that the genetic correlation between NLW and LAT is relatively high (rG = 0.714 ± 0.13), though could be different from 1 (*P* = 0.014). Here, we have shown that an LDPred2 model for PRS assuming infinitesimal causal loci drastically underperforms compared to one without this assumption (**Supplemental Figure 1, Supplemental Table 3**). Combining these two observations, we speculate that the disease architecture for ALL may be driven by a few large effect loci that are shared across ancestries. The lower genetic correlation between populations may then be driven by significant differences in the polygenic background, or by other yet-undiscovered population-enriched alleles. But as these loci may have smaller effects: PRS efficacy, and hence transferability, could be driven largely by the main effect loci, at least within the resolution of the sample size of the current validation cohort (i.e. CCLS). Future studies, particularly if focused on a single ethnic group such as NLW, that continue to elucidate the polygenic background of the ALL architecture may then both improve the accuracy of PRS model performance as well as exacerbating the loss of efficacy across populations that we are not currently able to detect. For this reason we would advocate for greater inclusion in GWAS representation despite currently observing little evidence in the loss of transferability in PRS model. With regards to the LAT population which has higher risk for ALL, increasing sample sizes will likely help improve PRS models in this population; indeed, using a smaller GWAS solely from LAT already substantially improved PRS prediction efficacy in out-of-sample LAT cohort (**Figure 1**) and further discovery of LAT-enriched alleles will improve PRS models for this population. More generally, diverse ancestries in multi-ancestry GWAS can also help with better fine-mapping of causal loci, which would improve both efficacy and transferability of PRS models.^15,17^

Altogether, our study provided guidance for future designs to propose and evaluate PRS models for ALL. Firstly, efforts to continue to increase the sample size and ancestry representation in GWAS is imperative. We have shown that genomic PRS using LDPred2 outperforms that based on just genome-wide significant loci (using the same GWAS, *i*.*e*. P+T models with P-value threshold of 5e-8). But this model currently does not outperform one simply based on aggregating all known loci from the literature, effectively combining information across multiple independent GWAS datasets. Therefore, an aggregation of available GWAS through a consortium effort should provide the ideal dataset to train better genomic PRS models. Efforts like the Childhood Leukemia International Consortium (CLIC)^28^ should provide the best resources in the foreseeable future. CLIC meta- or mega-analysis will include around 20,000 cases and 160,000 multi-ancestry ALL cases and controls. Given a larger dataset, we can ensure more homogeneity and greater sample size for both the testing and validation dataset and would have more cohorts with diverse ancestry to iteratively assess the transferability of the PRS models. This is particularly important for Latino populations, given the known heterogeneity in ancestry compositions and fine-scale structure of Latinos across the United States and Latin America.^29,30^

Secondly, given the suspected oligo-genic architecture of ALL, alternative PRS strategies that incorporate information from the distribution of effect sizes may also further improve the performance from a methodological standpoint. While LDPred2 controls somewhat the proportion of the genome underlying a trait through optimization of the ρ parameter, its prior is ultimately a “spike-and-slab” prior. A more direct modelling of the distribution of effect sizes, on top of a polygenic background may prove to be a better model for ALL. Methods following these types of models are emerging (e.g. see ref ^31^), and will likely become more mature in the near future. But even without a unified framework to model effect size distributions, a simpler approach^32^ that combines weighted PRS could also be more effective. In this case, one score would be derived from sections of the genome known to be associated with ALL that may also include multiple secondary but independent causal variants, and the other score could be derived from LDPred2 or similar approaches from the rest of the genome. The weights between these two scores can then be optimized in the training dataset as an additional parameter to derive a score that may outperform any of the existing models evaluated here.

PRS are intended to be robust prediction tools that would be utilized in research and clinical settings. In research settings, PRS would be applicable in defining the attributable fraction of leukemia risk derived from common genetic variation when examining other risks – either from low frequency genetic alleles or environmental factors. In the clinical setting, PRS could become useful tools to genetic counselors in working with children’s families in combination with sequencing for pathogenic germline variants in predisposition genes.

Ultimately, PRS may be incorporated with additional risk prediction tools such as markers of early leukemia-promoting mutations on a population scale in neonatal screening efforts where interventions were available. Employment of PRS in such settings requires accurate tools across all ancestral/ethnic groups, particularly for the Latino population who harbor the greatest risk of ALL, and our study represents one of the first approach toward this goal.

## MATERIALS AND METHODS

### Study Cohort

The California Childhood Cancer Record Linkage Project (CCRLP) includes all children born in California during 1982-2009 and diagnosed with ALL at the age of 0-14 years per California Cancer Registry records from 1988-2011. Children who were born in California during the same period and not reported to California Cancer Registry as having any childhood cancer were considered potential controls. Detailed information on sample matching, preparation and genotyping has been previously described^4^. Because ALL is a rare childhood cancer, to increase statistical power of a genetic study we followed previous practice^4^ and incorporated additional controls using adult individuals from the Kaiser Resource for Genetic Epidemiology Research on Aging Cohort (GERA; dbGaP accession: phs000788.v1.p2). The GERA cohort was chosen because a very similar genotyping platform had been used. Genome-wide SNP genotyping was performed for all individuals in CCRLP and GERA using the Affymetrix Axiom World LAT array^4^. Both studies included data on self-reported race/ethnicity. The imputation and quality control (QC) of SNP array data were carried out in each study population, as previously described in a multi-ancestry meta-analysis GWAS of ALL^2^. After QC filtering, the LAT GWAS included 1,878 cases and 8,441 controls, the NLW GWAS included 1,162 cases and 57,341 controls, the African American GWAS included 124 cases and 2067, and the East Asian GWAS included 318 cases and 5,017 controls.

Another GWAS was performed with individuals from the Children’s Oncology Group (COG; dbGAP accession: phs000638.v1.p1) as cases and from the Wellcome Trust Case– Control Consortium (WTCCC) as controls.^33^ We generally followed the same quality control pipeline, but because self-reported race/ethnicity was not available to us, we performed global ancestry estimations using ADMIXTURE and the 1000 Genomes populations as reference and removed individuals with < 90% estimated European ancestry from the analysis, resulting in a total of 1504 and 2931 NLW cases and controls respectively. This dataset was previously used as a replication cohort of European ancestry in our earlier study^2^, but here we combined it with CCRLP NLW to increase the sample size of the discovery GWAS (below).

The California Childhood Leukemia Study (CCLS)^12,34^, a non-overlapping California case-control study with controls selected from California birth records (1995-2008), was used as our validation dataset. In total, there were 306 NLW cases, 258 NLW controls, 592 LAT cases, and 509 LAT controls available for analysis. The QC procedures and imputation were performed in accordance with the discovery/training dataset.

### Overall Study Design

A PRS of an individual j is defined as a weighted sum of SNP allele counts:

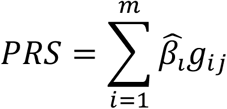

where m is the number of SNPs to be included in the predictor, 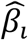 in the per allele weight for each SNP, *g*_*ij*_ is the allele count (0,1,2) or dosage of the allele of SNP in individual j.

For each step of score derivation, optimization, and evaluation, we used three non-overlapping datasets to (1) perform discovery GWAS to estimate variant effect sizes, (2) optimize parameters for the best predictive score, and (3) evaluate the predictive performance of the resulting scores (**Figure 1**). Following the convention previously suggested,^25,26^ we refer to the datasets used in each of the three steps as “GWAS”, “testing”, and “validation” datasets.

We randomly selected and held out 360 cases and 1,200 controls from each of CCRLP NLW (∼2.7% of the sample size) and LAT (∼15.2% of the sample size) as the testing datasets to identify the best PRS models, and used the remaining sample from CCRLP+GERA cohort as the GWAS dataset in the three different discovery GWAS: (1) NLW-only meta-analysis (combined with COG+WTCCC sample), (2) LAT-only GWAS, (3) multi-ancestry meta-analysis. For each GWAS, we constructed PRS using two established approaches - Pruning and Thresholding (P+T) and LDPred2.

### Discovery GWAS

We used PLINK (version 2.3 alpha) to test the association between imputed genotype dosage at each SNP and case-control status in logistic regression, after adjusting for the top 20 principal components to control for potential confounding due to fine-scale structure and variation in genetic ancestry within each ethnic group. For NLW and multi-ancestry GWAS meta-analysis, the results from each study and/or racial/ethnic groups were combined via the fixed-effect meta-analysis with variance weighting using METAL^35^. After excluding 360 cases and 1200 controls each for NLW and LAT (as the held-out/testing samples), we included 802 cases and 56141 controls in NLW, 1518 cases and 7210 controls in LAT, 318 cases and 5017 controls in East Asian, and 124 cases and 2067 controls in African American discovery GWAS in CCRLP/GERA. For NLW meta-analysis, CCRLP/GERA GWAS was meta-analyzed with a separate GWAS conducted with 1504 cases and 2931 controls from COG/WTCCC cohort, for a total sample size of 2306 cases and 59072 controls. Multi-ethnic meta-analysis was conducted with CCRLP/GERA NLW, LAT, East Asian, African American and COG/WTCCC individuals, totaling 4266 cases and 73366 controls. The total sample size for each discovery GWAS design can be found in **Supplemental Table 1**.

### Polygenic Risk Score Derivation / Optimization

For each ethnic-specific or multi-ancestry GWAS, we constructed PRS using two different methods: Pruning and Thresholding (P + T) and LDPred2. Both methods used the GWAS summary statistics as the starting point, but each make different choices for which SNPs to include in the predictor and the weight values assigned to each SNP.

*<u>Pruning and Thresholding (P+T)</u>* uses a P-value threshold and LD-driven clumping procedure to construct scores. The scores using P+T approach were constructed using PLINK (version 1.9). In brief, given a user-defined threshold for associated P-value and clumping parameters, the algorithm forms clumps around the index SNPs with all SNPs within a specified distance (kb) that have P-value and pair-wise LD (measured by r2) at levels greater than a specified threshold. The algorithm greedily and iteratively cycles through all index SNPs, beginning with the SNP with the most significant P-value, only allowing each SNP to appear in one clump. The most significant SNPs for each LD-based clump across the genome are used to build the PRS with associated estimate beta as weights. We constructed PRS using a range of P-values (1.0, 0.5, 0.05, 5 × 10^−4^, 5 × 10^−6^, and 5 × 10^−8^), r2 (0.2, 0.4, 0.6, and 0.8), and kb (250, 500) thresholds for a total of 48 PRS models to optimize under this approach.

*<u>LDPred2</u>* uses a Bayesian approach to calculate posterior mean effect size for each variant given a prior and subsequent shrinkage based on the extent to which the variant is correlated with similarly associated variants.^27,36^ The underlying Gaussian distribution additionally considers the proportion of causal variants (ρ). LDPred2 uses a grid of values for hyper-parameters/tuning parameter - ρ, *h*^2^ (the SNP heritability), and sparsity (whether to fit some variant effects to exactly zero) to construct PRS. We used ρ from a sequence of 17 values from 10^−4^ to 1 on a log-scale, a range of *h*^2^ within (0.7, 1, 1.4) × estimated heritability, and a binary sparsity option of either on and off (LDPred2-grid models). In addition, we tested a model assuming infinitesimal causal effects, where each variant assumed to contribute to disease risk (LDPred2-inf model). In total, we evaluated 103 PRS models using LDPred2.

Once the variants and weights for each PRS model were estimated, the scores were generated in the testing sample (360 cases and 1200 controls in NLW or LAT) using PLINK (version 2.3 alpha), and then standardized to have mean of 0 and variance of 1. For each strategy, the score with the best predictive performance was determined based on the highest Negelkerke’s pseudo-R^2^, which was calculated as the difference of R^2^ from a full model inclusive of the PRS and the covariates and the R^2^ from a null model with covariates alone. Covariates in the model included the first 20 principal components (PCs) and sex.

### PRS evaluation

After optimizing the PRS model in held-out testing samples of 360 cases and 1200 controls, we computed the PRS score in the CCLS, which is our validation dataset. The CCLS included 306 cases and 258 controls in the NLW subcohort, and 592 cases and 509 controls in the LAT subcohort. The predictive performance of PRS was quantified by Negelkerke’s pseudo-R^2^ (the proportion of variance explained, as computed above for the testing datasets) and area under receiver operating characteristic curve (AUC; probability that a case ranks higher than a control). AUC was computed for the full model with covariates to account for population stratification. AUC for the null model (ALL ∼ 10 PCs + sex) is 0.593 and 0.577 in CCLS LAT and NLW, respectively. AUC were calculated using pROC package in R.^37^ Standard errors in these measures of model performance was computed with 1,000 sets of bootstrap samples across individuals.

### Ancestry Inference

Global ancestry inference was performed on CCRLP Latino cases and controls using RFMix^38^, using a reference panel consisting of 671 non-Finnish European individuals for European ancestry, 716 African individuals for African ancestry, and 94 Admixed American individuals (7 Columbian, 12 Karitianan, 14 Mayan, 4 Mexican in Los Angeles, 37 Peruvian in Lima, Peru, 12 Pima, and 8 Surui) for Indigenous American (IA) ancestry from gnomAD v3.1 release^39^, as previously identified to be enriched with indigenous ancestry.^40^ We stratified Latino individuals into three tertiles of global European ancestry, and in each group evaluated the predictive performance of the best PRS model for NLW_NLW strategy (*i*.*e*. GWAS conducted in NLW, and PRS optimization performed in held-out NLW).

## Data Availability

Our data is derived from the California Biobank. We respectfully are unable to share raw, individual genetic data freely with other investigators since the samples and the data are the property of the State of California. Should we be contacted by other investigators who would like to use the data, we will direct them to the California Department of Public Health Institutional Review Board to establish their own approved protocol to utilize the data, which can then be shared peer-to-peer. The State has provided guidance on data sharing noted in the statement below: "California has determined that researchers requesting the use of California Biobank biospecimens for their studies will need to seek an exemption from NIH or other granting or funder requirements regarding the uploading of study results into an external bank or repository (including into the NIH dbGaP or other bank or repository). This applies to any uploading of genomic data and/or sharing of these biospecimens or individual data derived from these biospecimens. Such activities have been determined to violate the statutory scheme at California Health and Safety Code Section 124980 (j), 124991 (b), (g), (h) and 103850 (a) and (d), which protect the confidential nature of biospecimens and individual data derived from biospecimens. Investigators may agree to share aggregate data on SNP frequency and their associated p-values with other investigators and may upload such frequencies into repositories including the NIH dbGaP repository providing: a) the denominator from which the data is derived includes no fewer than 20,000 individuals; b) no cell count is for < 5 individuals; and c) no correlations or linkage probabilities between SNPs are provided.) Since our dataset is derived from less than 20,000 subjects, we are not able to upload the data to dbGAP or another repository. All underlying numerical data used to create figures are available at https://doi.org/10.7910/DVN/FFBYRT.
All other datasets not derived from the California Biobank are available on dbGAP.

## ACKNOWLEDGEMENTS

This work was supported by research grants from the National Institutes of Health (R01CA155461, R01CA175737, R01ES009137, P42ES004705, P01ES018172, P42ES0470518, R24ES028524, and R01CA262263) and the Environmental Protection Agency (RD83451101), United States. C.W.K.C. is supported by R35GM142783 from the Natinoal Institute of General Medical Sciences (NIGMS). The content is solely the responsibility of the authors and does not necessarily represent the official views of the National Institutes of Health and the EPA. The collection of cancer incidence data used in this study was supported by the California Department of Public Health as part of the statewide cancer reporting program mandated by California Health and Safety Code Section 103885; the National Cancer Institute’s Surveillance, Epidemiology and End Results Program under contract HHSN261201000140C awarded to the Cancer Prevention Institute of California, contract HHSN261201000035C awarded to the University of Southern California, and contract HHSN261201000034C awarded to the Public Health Institute; and the Centers for Disease Control and Prevention’s National Program of Cancer Registries, under agreement U58DP003862-01 awarded to the California Department of Public Health. The biospecimens and/or data used in this study were obtained from the California Biobank Program, (SIS request #26), Section 6555(b), 17 CCR. The California Department of Public Health is not responsible for the results or conclusions drawn by the authors of this publication. This study makes use of data generated by the Wellcome Trust Case– Control Consortium. A full list of the investigators who contributed to the generation of the data is available from www.wtccc.org.uk. Funding for the project was provided by the Wellcome Trust under award 076113 and 085475. For recruitment of subjects enrolled in the CCLS replication set, the authors gratefully acknowledge the clinical investigators at the following collaborating hospitals: University of California Davis Medical Center (Dr. Jonathan Ducore), University of California San Francisco (Drs. Mignon Loh and Katherine Matthay), Children’s Hospital of Central California (Dr. Vonda Crouse), Lucile Packard Children’s Hospital (Dr. Gary Dahl), Children’s Hospital Oakland (Dr. James Feusner), Kaiser Permanente Roseville (formerly Sacramento) (Drs. Kent Jolly and Vincent Kiley), Kaiser Permanente Santa Clara (Drs. Carolyn Russo, Alan Wong, and Denah Taggart), Kaiser Permanente San Francisco (Dr. Kenneth Leung), and Kaiser Permanente Oakland (Drs. Daniel Kronish and Stacy Month). The authors additionally thank the families for their participation in the California Childhood Leukemia Study (formerly known as the Northern California Childhood Leukemia Study). Finally, the authors acknowledge the Center for Advanced Research Computing (CARC; https://carc.usc.edu) at the University of Southern California for providing computing resources that have contributed to the research results reported within this publication.

