## Supplemental Figures and Tables for "Evaluating Genomic Polygenic Risk Scores for Childhood Acute Lymphoblastic Leukemia in Latinos"

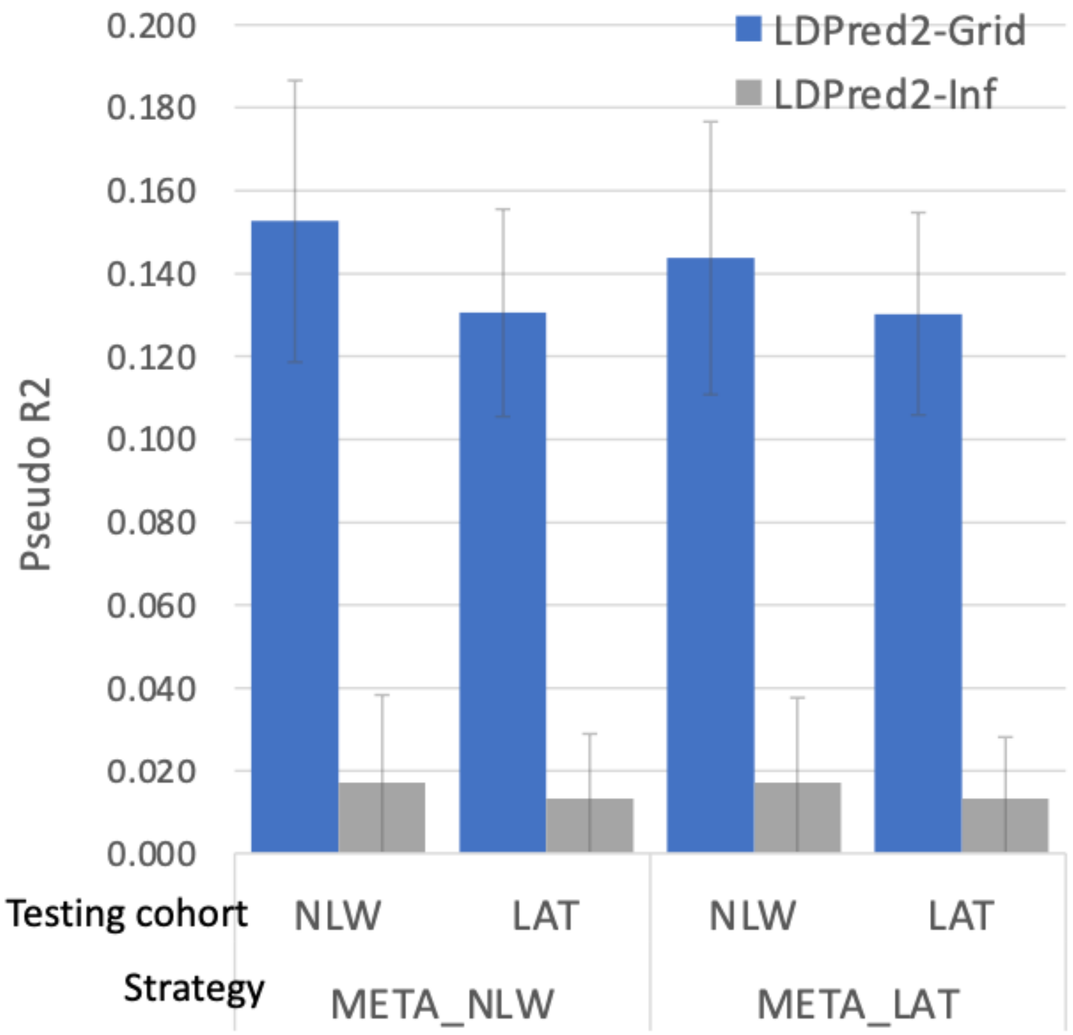


**Supplemental Figure 1: Pseudo $R^2$ of best performing LDPred2-Grid model vs. LDPred2-Inf in META_NLW and META_LAT strategies.** These models were tested in CCLS NLW or LAT. In all cases, the LDPred2-Grid models are substantially better than LDPred2-Inf models that assumes an infinitesimal genetic architecture.

| Strategy | Discovery GWAS | Testing dataset |
|---|---|---|
| NLW_NLW | 2306 case; 59072 control NLW | 360 case; 1200 control NLW |
| NLW_LAT | 2306 case; 59072 control NLW | 360 case; 1200 control LAT |
| META_NLW | 4266 case; 73366 control multi-ancestry | 360 case; 1200 control NLW |
| META_LAT | 4266 case; 73366 control multi-ancestry | 360 case; 1200 control LAT |
| LAT_LAT | 1518 case; 7210 control LAT | 360 case; 1200 control LAT |

**Supplemental Table 1: Summary of sample size for each strategy.** Strategies follow the $POP_{GWAS}\_POP_{testing}$ convention, where the first population is used to conduct the discovery GWAS, and the second population is used to derive and optimize the PRS model. NLW denotes non-Latino Whites; LAT denotes Latinos; META denotes multi-ancestry meta-analysis

| Strategy | Approach | Parameters | SNP count | P-value | AUC | SE_AUC | PseudoR2 | SE_PseudoR2 |
|---|---|---|---|---|---|---|---|---|
| NLW_NLW | LDPred2 | rho=0.0032 h2=0.2165 nosparse | 1078940 | 3.95E-12 | 0.652 | 0.032 | 0.060 | 0.020 |
| NLW_LAT | LDPred2 | rho=0.01 h2=0.1826 sparse | 1083465 | 7.30E-09 | 0.635 | 0.033 | 0.041 | 0.018 |
| META_NLW | LDPred2 | rho=0.0032 h2=0.1376 sparse | 1078940 | 8.70E-24 | 0.700 | 0.031 | 0.131 | 0.025 |
| META_LAT | LDPred2 | rho=0.001 h2=0.1127 nosparse | 1083365 | 1.28E-23 | 0.701 | 0.031 | 0.130 | 0.024 |
| LAT_LAT | LDPred2 | rho=0.001 h2=0.1764 sparse | 1083365 | 1.75E-21 | 0.699 | 0.031 | 0.116 | 0.026 |

**Supplemental Table 2: Summary of best performing model for each strategy tested on CCLS LAT.** The best PRS model under each strategy to construct PRS is selected out of two separate approaches (Pruning-and-thresholding and LDPred2) based on PseudoR2. The parameters of the best model in each strategy is given. For LDPred2, rho is the proportion of causal variants. h2 is the estimated heritability. Sparse or no sparse denote the inference mode where some variant effects are fit to exactly zero. SNP count is the number of SNPs retained in each of the best PRS model. P-value denotes the evidence of association of the PRS model in a logistic regression model with additional covariates of 20 PCs and sex. AUC denotes area under the curve from receiveer-operator characteristic analysis. PseudoR2 was computed as the difference between a logistic regression model with PRS and one without PRS. SE denotes standard error for both AUC and PseudoR2, which were computed using 1,000 bootstrap samples.

| Strategy | approach | CCLS NLW PseudoR2 | CCLS NLW SE | CCLS NLW AUC | CCLS LAT PseudoR2 | CCLS LAT SE | CCLS LAT AUC |
|---|---|---|---|---|---|---|---|
| META_NLW | LDPred2-Grid | 0.153 | 0.034 | 0.710 | 0.131 | 0.025 | 0.700 |
| | LDPred2-Inf | 0.017 | 0.021 | 0.591 | 0.013 | 0.016 | 0.609 |
| META_LAT | LDPred2-Grid | 0.144 | 0.033 | 0.704 | 0.130 | 0.024 | 0.701 |
| | LDPred2-Inf | 0.017 | 0.021 | 0.590 | 0.013 | 0.015 | 0.609 |

**Supplemental Table 3: Pseudo R2 of best performing LDPred2-Grid model vs. LDPred2-Inf.** LDPred2-Inf is a LDPred2 model that assumes the infinitesimal model. P-value denotes the evidence of association of the PRS model in a logistic regression model with additional covariates of 20 PCs and sex. AUC denotes area under the curve from receiveer-operator characteristic analysis. PseudoR2 was computed as the difference between a logistic regression model with PRS and one without PRS. SE denotes standard error for both AUC and PseudoR2, which are computed using 1,000 bootstrap samples.

| Gene | SNP | Effect Allele | beta |
|---|---|---|---|
| C5orf56 | 5:131811182:G:A | G | 0.1207 |
| BAK1 | 6:33546837:T:C | C | 0.1813 |
| MYB | 6:135411228:T:C | T | 0.2425 |
| IKZF1 | 7:50459043:G:A | A | 0.4897* |
| IKZF1 | 7:50477144:G:A | A | 0.3678 |
| 8q24 | 8:130156143:A:G | G | 0.2536 |
| CDKN2A | 9:21975319:T:A | A | 0.6307 |
| CDKN2A | 9:21993964:T:C | T | 0.3037* |
| TLE1 | 9:83728588:C:T | C | 0.2226 |
| GATA3 | 10:8104208:C:A | A | 0.19 |
| BMI1 | 10:22374489:G:A | G | 0.2231 |
| PIP4K2A | 10:22853102:T:C | C | 0.2772 |
| ARID5B | 10:63721176:C:T | C | 0.4959 |
| JMJD1C | 10:65020890:A:G | A | 0.1846 |
| TET1 | 10:70329064:G:A | A | 0.2086 |
| LHPP | 10:126293309:A:G | G | 0.1103 |
| ELK3 | 12:96645605:A:C | C | 0.2053 |
| CEBPE | 14:23592617:A:C | A | 0.1768* |
| CEBPE | 14:23589349:A:G | G | 0.3322 |
| IKZF3 | 17:37957235:T:C | C | 0.7456 |
| IKZF3 | 17:37983492:T:C | T | 0.1857* |
| IGF2BP1 | 17:47217004:C:G | C | 0.107 |
| ERG | 21:39784752:T:C | C | 0.1353 |

**Supplemental Table 4: List of SNPs used to construct PRS based on previously known risk loci.** Gene names (gene) are given based on the nearest gene unless the variant is in gene desert. SNP positions are given in hg19 coordinates. Effect sizes (beta) is from previous meta-analysis (Jeon et al. Leukemia 2022). * denotes that the effect size used was from conditional analysis as these are secondary signals in the locus.
